# Power Law in COVID-19 Cases in China

**DOI:** 10.1101/2020.07.25.20161984

**Authors:** Behzod B. Ahundjanov, Sherzod B. Akhundjanov, Botir B. Okhunjanov

## Abstract

The novel coronavirus (COVID-19) was first identified in China in December 2019. Within a short period of time, the infectious disease has spread far and wide. This study focuses on the distribution of COVID-19 confirmed cases in China—the original epicenter of the outbreak. We show that the upper tail of COVID-19 cases in Chinese cities is well described by a power law distribution, with exponent around one in the early phases of the outbreak (when the number of cases was growing rapidly) and less than one thereafter. This finding is significant because it implies that (i) COVID-19 cases in China is heavy-tailed and disperse; (ii) a few cities account for a disproportionate share of COVID-19 cases; and (iii) the distribution generally has no finite mean or variance. We find that a proportionate random growth model predicated by Gibrat’s law offers a plausible explanation for the emergence of a power law in the distribution of COVID-19 cases in Chinese cities in the early phases of the outbreak.

## 1 Introduction

The coronavirus disease 2019 (COVID-19) was first discovered in Wuhan region of China in December 2019 (Zhu et al., 2020). The contagious disease quickly spread within China, despite unprecedented and aggressive containment measures, and crossed the borders reaching every corner of the world within a short period of time, with the World Health Organization (WHO) declaring COVID-19 outbreak a global pandemic on March 11, 2020 (Cucinotta and Vanelli, 2020). This study focuses on the distribution of COVID-19 confirmed cases in China—the original epicenter of the outbreak—in the first wave of the epidemic. The presence of Chinese cities with very large number of COVID-19 cases, the very wide dispersion in COVID-19 cases across China, and the effect of the pandemic on the economy and welfare make it crucial for researchers and policymakers alike to better understand COVID-19 distribution for effective planning and policy design as well as more efficient use of government resources.

In this article, we demonstrate that the right tail of the distribution of COVID-19 confirmed cases in Chinese cities is well-characterized by a power law (Pareto) distribution, meaning that the probability that a number of COVID-19 cases is more than *x* is roughly proportional to *x*^−*γ*^, i.e., Prob(*X > x*) ∼ *x*^−*γ*^, where *γ* is the power law (Pareto) exponent.^1^ While the estimated power law exponent is *γ* ≃ 1 in the early phases of the outbreak, when the number of cases was rising precipitously, it is *γ <* 1 thereafter, which indicates the fitted power law distribution in general has no finite moments, including mean and variance. The power law fit is robust to a range of estimation methods and goodness-of-fit tests, and the distribution outperforms several alternative distributions in fitting the data. Power law distributions are characterized by heavy tails, which make the likelihood of extreme (upper-tail) events more typical. In case of COVID-19, this implies an extremely large number of cases becomes more likely, which is actually true for China, where a few cities had extremely large number of cases (Han et al., 2020).

Power laws are extraordinarily ubiquitous in the social and natural sciences, having been confirmed for the distributions of income and wealth (Pareto, 1896; Champernowne, 1953; Wold and Whittle, 1957; Singh and Maddala, 1976; Klass et al., 2006; Benhabib et al., 2011; Toda, 2012), consumption (Toda and Walsh, 2015; Toda, 2017), firm size (Stanley et al., 1995; Axtell, 2001; Luttmer, 2007), farmland (Akhundjanov and Chamberlain, 2019), city size (Krugman, 1996; Gabaix, 1999; Ioannides and Overman, 2003; Berliant and Watanabe, 2015; Devadoss et al., 2016), natural gas and oil production (Balthrop, 2016), carbon dioxide (CO_2_) emissions (Akhundjanov et al., 2017), forest fires (Malamud et al., 1998), earthquakes (Bak and Tang, 1989), frequency of words (Zipf, 1949; Irmay, 1997), and even university research activities (Plerou et al., 1999). The omnipresence of power laws is partly explained by the fact that they are preserved over an extensive array of mathematical transformations (Gabaix, 2009). In fact, any multiplicative transformation of this distribution will, in theory, be power-law distributed as well. This explains why a power-law distribution is also referred to as a scale-free distribution. Our paper contributes to this body of work by presenting evidence for the existence of power law in an epidemiological context.

An interesting aspect of power law distribution is that it is the macro-level steady-state phenomenon that, in theory, can arise from a micro-level proportionate random growth process, known as Gibrat’s law (Gibrat, 1931). According to Gibrat’s law of proportionate growth, each unit’s (e.g., city’s) growth rate is drawn randomly and independently of its size, meaning units (whether large or small) on average grow at similar rates.^2^ This empirical regularity, similar to power laws, has been documented extensively in the social and natural sciences.^3^ In this study, we formally test for proportionate random growth at micro-level by analyzing growth rates of COVID-19 cases in Chinese cities. We find that a proportionate random growth model is a plausible power-law generating mechanism for COVID-19 cases in China in the early phases of the outbreak, when the number of cases was growing. In the later phases of the outbreak, as the number of cumulative cases starts to stabilize, a proportionate random growth model becomes less plausible. This is expected as, after all, Gibrat’s law of proportionate growth is a model of growth. As with any theory, we remain cognizant of Gabaix‘s (2009) remark that, “the main question of empirical work should be how well a theory fits, rather than whether it fits perfectly.”

There are a couple of noteworthy implications of the findings presented in this article. First, and foremost, the analysis uncovers potential heavy-tailedness and tail risk property in COVID-19 cases in Chinese cities, which has a direct implication for empirical research. Specifically, it shows that thin-tailed distributions (e.g., the normal) are *not* appropriate for COVID-19 cases in Chinese cities as such distributions dismiss extremely large number of cases as an improbable observation. Our analysis reveals that even some, more common heavy-tailed distributions (e.g., the lognormal and exponential) are not plausible in this context. This is important because distributions like normal or lognormal are often ‘go-to’ distributions in empirical work, and the trend of adopting these distributions with little or no a priori justification or on the basis of convenience has been observed in the rapidly expanding COVID-19 literature. On the other hand, a power-law distribution is able to capture the heavy upper-tail of the data more closely, outperforming a number of alternative distributions. For sound analysis of the effects of COVID-19 pandemic, using statistically justified and robust methods that account for possible heavy-tailedness and tail risk properties is integral (Distaso et al., 2020).

Second, given the estimated Pareto exponent is generally less than one (*γ <* 1), the distribution of COVID-19 cases in Chinese cities is heavy-tailed and so disperse that observations near the mean account for little of the cumulative distribution of COVID-19 cases. This implies talking about the typical or average number of COVID-19 cases is inconsequential as it does not represent the majority of cases. In fact, even though it is possible to compute sample mean and variance for the observed data, these moments are generally non-convergent. Therefore, the distribution cannot be well characterized by quoting its mean and variance. Instead, quantile analysis or order statistics would be a more appropriate approach to describing the data.

Finally, the heavy upper-tail of the distribution is suggestive of concentration of COVID-19 cases in China, with the total cases essentially being determined by a few cities that bore the brunt of the outbreak, which is true in case of China (Han et al., 2020). This has implications for more effective epidemiological planning and policy design as by directing resources to disease epicenters (i.e., cities located in the upper tail), the spread of the outbreak can potentially be contained or, at least, slowed. Moreover, by understanding characteristics of these upper-tail cities that make them especially prone to the infection, more effective preventative measures can be designed and implemented to combat possible subsequent waves of the pandemic.

The literature in this area is thin, but gradually forming. In the concurrent work, Beare and Toda (2020), studying the distribution of COVID-19 confirmed cases for US counties, find that the upper tail of this distribution follows a power law, with Pareto exponent close to 1. Similarly, Blasius (2020), examining the distribution of COVID-19 confirmed cases and deaths for US counties, concludes that both distributions exhibit a power-law behavior. Our paper contributes to this nascent line of literature by exploring the distribution of COVID-19 confirmed cases in China—the origin of the outbreak. A distinctive feature of our study is that COVID-19 cases in China affords us to capture the entire life cycle of the pandemic (at least in its first wave): outbreak detection, spread, peak, and decline to zero new daily cases. In contrast, the analyses presented in Beare and Toda (2020) and Blasius (2020) are based on datasets that were largely evolving at the time, as both the United States and a whole host of other countries were battling to contain the spread of the virus when these articles were initially written. Thus, the results of the above studies are likely subject to change with newer data.

The remainder of the paper is structured as follows. Section 2 introduces the data for the analysis. Section 3 presents the methods and findings for power law analysis. Section 4 reviews a prominent mechanism that can generate power-law distribution, and discusses its plausibility in the case examined in this study. Section 5 provides some concluding remarks.

## 2 Data

Daily data on the cumulative number of COVID-19 confirmed cases for Chinese cities come from Harvard Dataverse (China Data Lab, 2020). The dataset includes 339 cities in China and tracks the COVID-19 cases starting from January 15, 2020. Our main analysis focuses on COVID-19 cases as of May 23, 2020, the latest data on cumulative cases at the time of writing this paper. By May 23, the number of cumulative cases in China had stabilized, with zero new daily cases, which suggests the containment of the first wave of the outbreak in the country. Our analysis thus allows us to understand the distribution of COVID-19 cases in its first complete life cycle.

Figure 1 shows the location of Chinese cities with confirmed COVID-19 cases as of May 23 on the map of China. As illustrated in Figure 2, most Chinese cities in the sample had reported a positive number of cases by February 8, 2020. Figure 3 shows the evolution of empirical distribution of COVID-19 cases in Chinese cities over select dates. It is apparent that the distribution has been right-skewed, with heavier right tail. Also, the distribution has been gradually sliding rightward over time, which reflects growing number of COVID-19 cases across Chinese cities.

**Figure 1:**
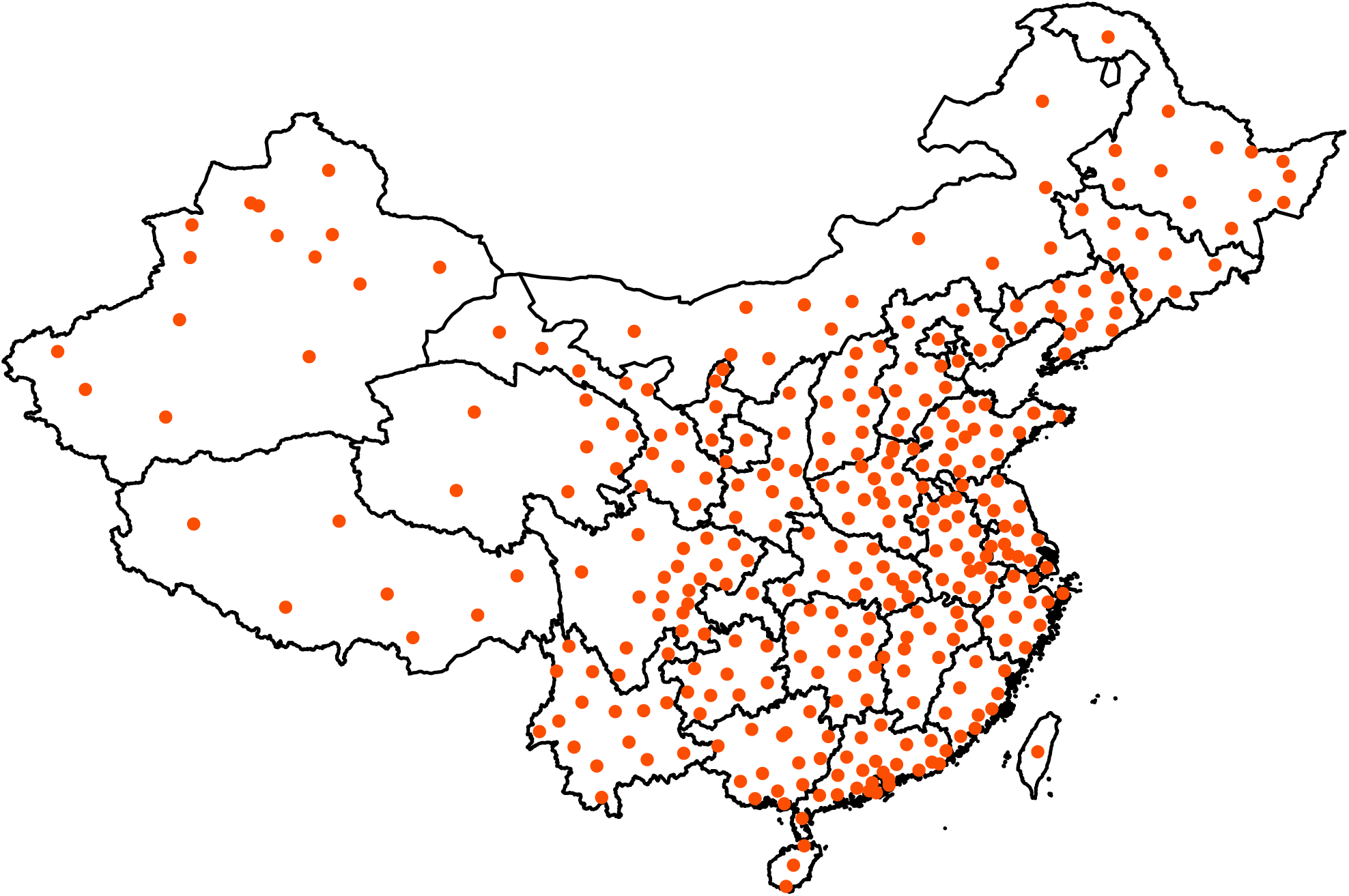
Chinese cities with confirmed COVID-19 cases as of May 23, 2020. Data source: Harvard Dataverse (China Data Lab, 2020).

**Figure 2:**
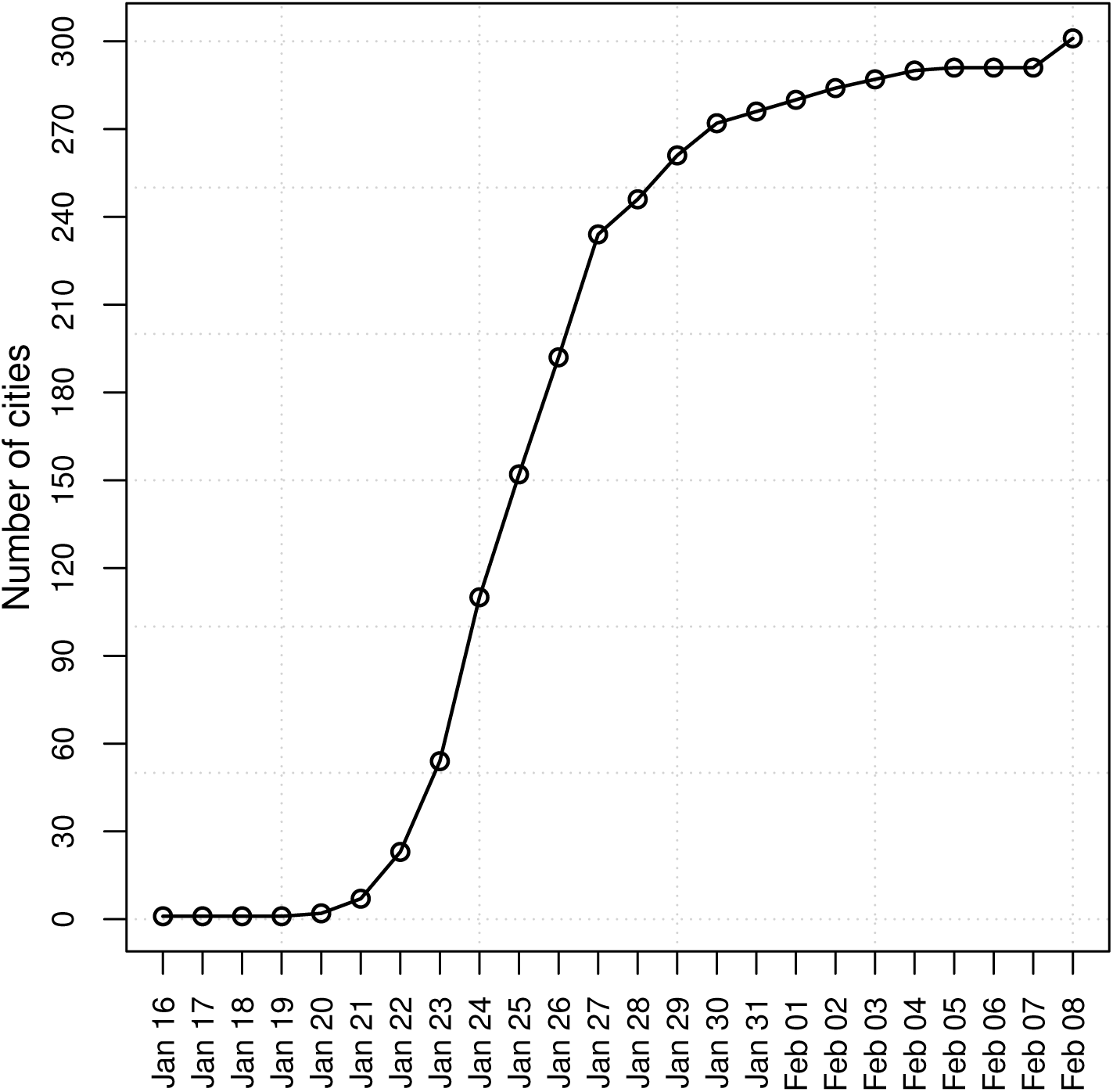
The number of Chinese cities with confirmed COVID-19 cases over time. Data source: Harvard Dataverse (China Data Lab, 2020).

**Figure 3:**
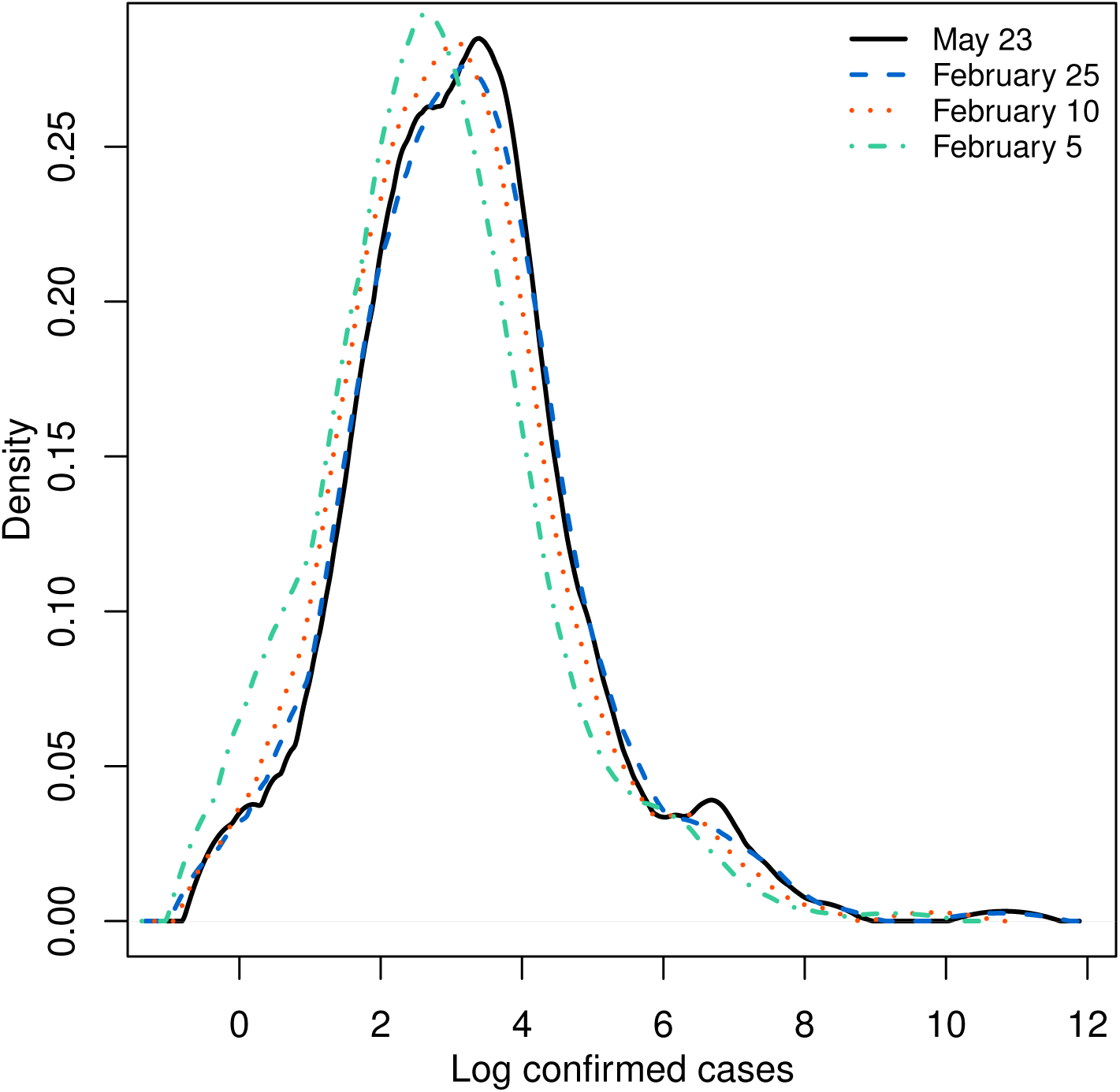
The empirical distribution of cumulative number of COVID-19 confirmed cases for Chinese cities. The empirical distribution is obtained using kernel density with Epanechnikov kernel and the smoothing band-width based on unbiased cross-validation method.

In Figure 4, we plot the ratio of COVID-19 cases to population size in Chinese cities between January 15, 2020 and May 23, 2020.^4^ It is evident that COVID-19 cases represented a minute fraction of total population throughout the study period, with majority of cities having less than 0.00025 fraction of their population infected by the virus. Even Wuhan, a city pummeled by the outbreak, had only 0.006 fraction of its population with confirmed COVID-19 infection by the end of the first wave of the pandemic. This shows that the population of cities in the beginning, and throughout the first wave, of the epidemic does not limit its spread, meaning the population of cities can effectively be treated as “infinite” relative to COVID-19 cases.

**Figure 4:**
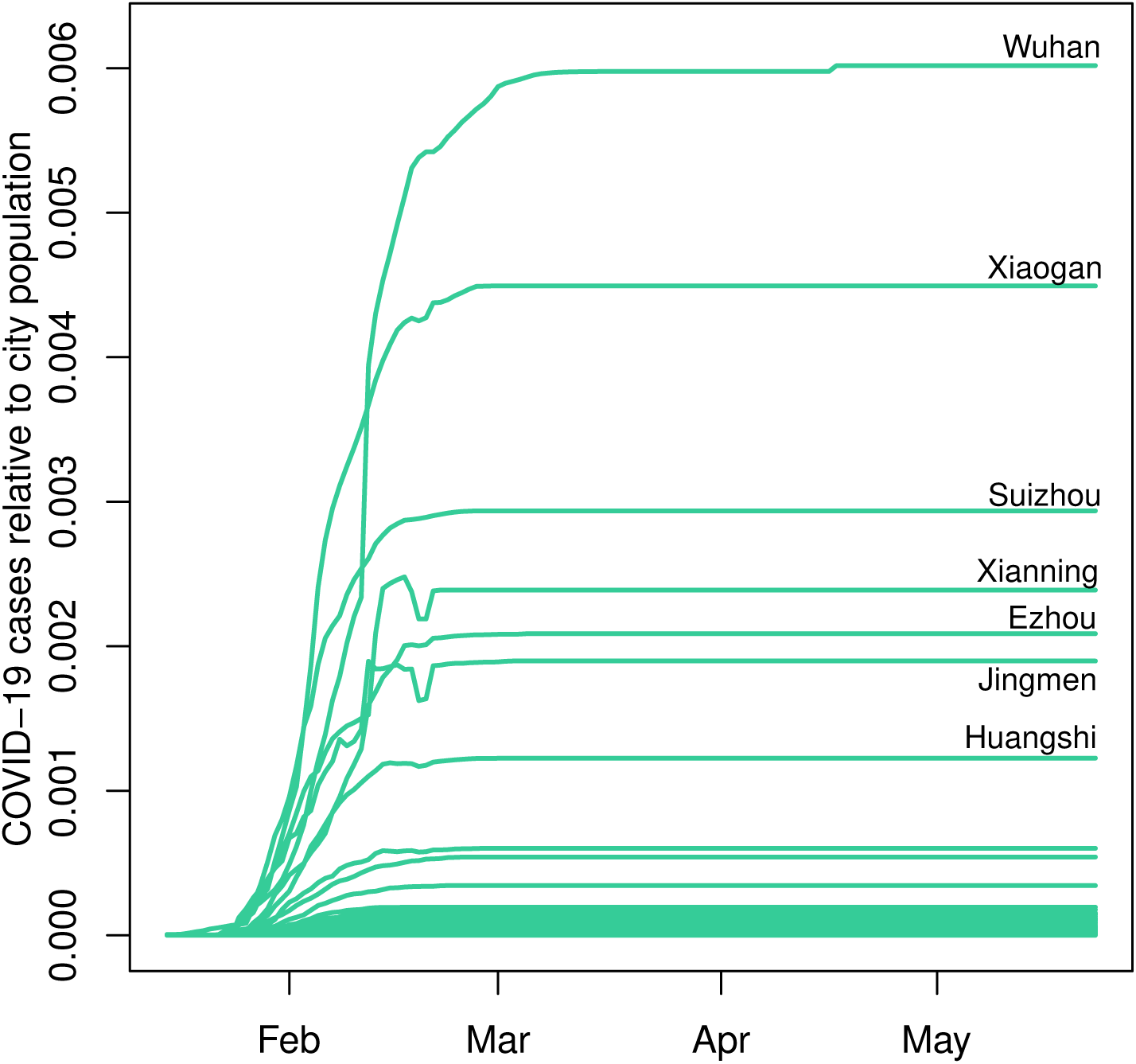
The ratio of COVID-19 cases to city population in China between January 15, 2020 and May 23, 2020.

A power law analysis is data intensive, with Clauset et al. (2009) recommending a minimum of 50 observations for reliable analysis. This condition is well-satisfied here, including for the upper tail of our sample, as discussed further below.

## 3 Power law analysis

In this section, we examine the distribution of the cumulative number of COVID-19 confirmed cases for Chinese cities. We first present the methodology for power law analysis, followed by estimation results and diagnostics.

### 3.1 Power-law parameter estimation

Suppose *X* is a random variable whose data generating process is a continuous power law (Pareto) distribution. The corresponding probability distribution function (PDF) is specified as

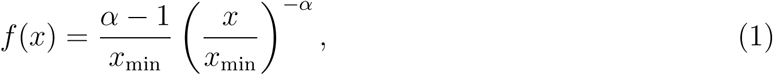

where *x* is an outcome of *X* for *x* ∈ ℝ_+_, where ℝ_+_ = {*x* ∈ ℝ|*x >* 0}, *x*_min_ is the threshold beyond which (i.e., *x* ≥ *x*_min_) power-law behavior sets in, and *α* is the power-law (Pareto) exponent, which is a parameter of interest. The *m*th non-central moment for the power law distribution is given by

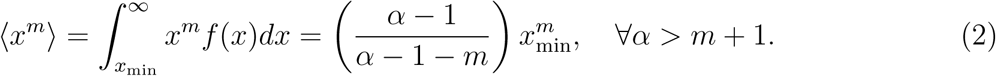

Hence only the first ⌊*α* − 1⌋ moments exist for *m < α* − 1. Although higher-order moments can be calculated for any finite sample, these estimates do not asymptotically converge to any particular value. Given the sample *x*_1_, …, *x*_*n*_, the joint log-likelihood function can be written as

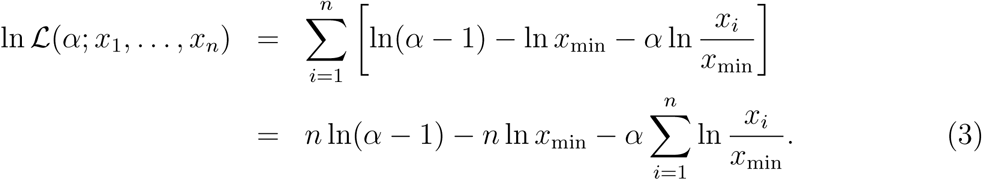

First-order condition yields the maximum likelihood estimate (MLE) of

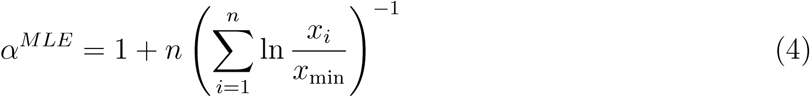

with the standard error (SE) of the estimate given by

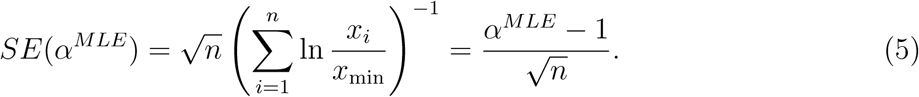

It is standard to report the counter-cumulative parameter *γ* = *α*^*MLE*^ − 1, known as the Hill estimator (Hill, 1975), instead of (4). The Hill estimator, which is obtained from (4) after a small-sample adjustment, takes the following form

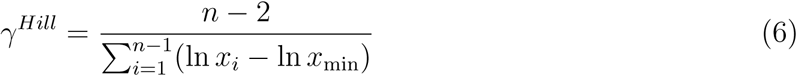

with the standard error of the estimate given by

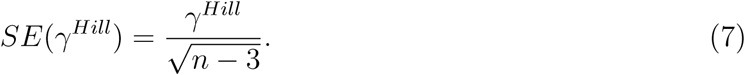

The power-law fit to data is typically illustrated by plotting the counter-cumulative distribution function (counter-CDF)—also known as the survival function—on doubly logarithmic axes. The counter-CDF of a power law is specified as

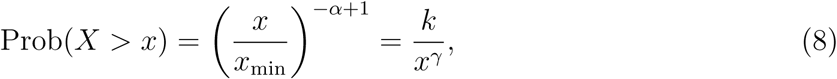

where 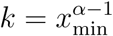 is a constant. Taking the log of both sides of (8) yields a well-known linear relationship between log counter-cumulative probability (i.e., ln Prob(*X > x*)) and log data (i.e., ln *x*), with the counter-cumulative parameter −*γ* being the slope of the line.

An alternative approach to estimating the counter-cumulative parameter *γ* is through a regression-based technique. Specifically, estimate the following regression equation with ordinary least squares (OLS)

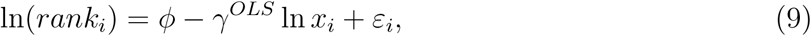

where *rank*_*i*_ is observation *i*’s rank in the distribution, *ϕ* is the intercept term, *γ*^*OLS*^ is the parameter of interest, and *ε*_*i*_ is the idiosyncratic disturbance term. Equation (9) also shows that a power law distributed process appears approximately linear on a log-log plot of *rank*_*i*_ against *x*_*i*_, with slope of −*γ*^*OLS*^. The asymptotic standard error for *γ*^*OLS*^ is given by (Gabaix and Ibragimov, 2011)

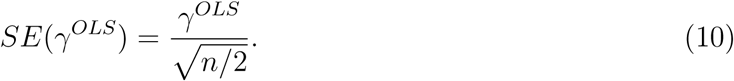

An important consideration in power law analysis is the specification of the threshold parameter *x*_min_, beyond which power-law behavior takes hold. There are several approaches proposed in the literature in this regard. For instance, one strand of literature suggests to select *x*_min_ at either the 95% quantile of the data or the point where empirical PDF or CDF roughly straightens out on a log-log plot (Gabaix, 2009). Another strand of the literature simply uses the largest 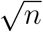 or *n*/10 observations in the analysis (Farmer et al., 2004). Clearly, these approaches are rather arbitrary and thus suffer from a certain degree of uncertainty about whether they can capture the true starting point of power-law behavior. If the arbitrarily selected *x*_min_ is smaller than the true value of *x*_min_, the estimate of power-law exponent will be biased as it attempts to fit non-power law data to a power law model. Conversely, if *x*_min_ exceeds the point where the true power-law behavior begins, it results in discarding valuable data and causes the standard error to increase. Importantly, Perline (2005), investigating the empirical consequences of this concern, shows that sufficiently truncated Gumbel-type distributions (e.g., the lognormal) can also produce a linear pattern on a log-log plot, hence imitating the power law distribution.

We adopt a more systematic, data-driven procedure to select *x*_min_ (Clauset et al., 2009). The main advantage of this method is that it allows the analyst to identify the power-law portion of the data (if any) in a more objective and principled manner, balancing the tradeoff between setting *x*_min_ too small or too high relative to the true value of *x*_min_. This approach essentially treats each observation in the sample as a potential candidate for *x*_min_ and selects the best candidate based on the minimization of the Kolmogorov-Smirnov (KS) goodness-of-fit statistic, which is given by

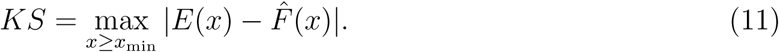

In (11), *E*(*x*) is the empirical CDF and 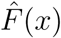 is the estimated power-law CDF. The optimal *x*_min_ minimizes the distance between the empirical CDF and the estimated power-law CDF. The computational algorithm takes the following form:

Step 1: Set *x*_min_ = *x*_1_;

Step 2: Perform power-law parameter estimation using *x* ≥ *x*_min_;

Step 3: Compute the KS statistic in (11);

Step 4: Repeat steps 1-3 for all *x*_*i*_ for *i* = 1, …, *n*; Step 5: Select *x*_min_ with the lowest KS statistic.

### 3.2 The goodness-of-fit tests

Significant parameter estimates alone do not provide sufficient evidence in favor of power-law fit to data. Power-law analysis is accompanied by a series of diagnostic tests. To guard against potential misspecification issues, one needs to conduct a goodness-of-fit test and compare the power-law fit to data with those of alternative distributions.

Gabaix and Ibragimov (2011) proposed ‘rank – 1/2’ test to verify the goodness-of-fit of power law distribution. Let *x*^*^ be defined as

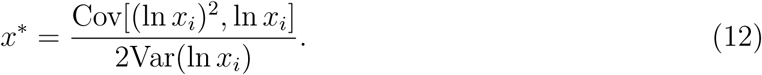

Then, regress bias-adjusted log rank against the log data and a quadratic deviation term, as in

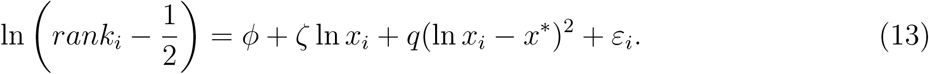

The goodness-of-fit statistic is specified as *q/ζ*^2^. The null hypothesis of power-law distributedness is rejected if *q/ζ*^2^ *>* 1.95(2*n*)^−1*/*2^, where the latter term is the goodness-of-fit threshold.

Further, Clauset et al. (2009) suggest comparing power-law fit with those of other competing (heavy-tailed) distributions, such as the lognormal and exponential. Accordingly, we fit these alternative distributions to the data by MLE and provide visual comparisons of the distributions’ fits on a doubly logarithmic plot as detailed above. In addition to conventional model fit measures, such as the Akaike information criterion (AIC) and Bayesian information criterion (BIC), we also implement the likelihood ratio test of Clauset et al. (2009) for a formal comparison. The likelihood ratio statistic is specified as

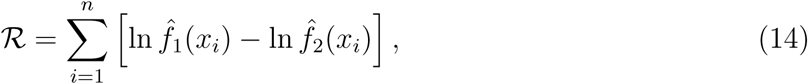

where 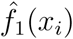 and 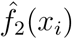 are the probabilities predicted by power law and an alternative distribution, respectively. If the likelihood ratio statistic is positive, it indicates the power law distribution fits the data more closely. If it is negative, then an alternative distribution yields a better fit.^5^

### 3.3 Application

The methods discussed in Sections 3.1 and 3.2 are applied to the cumulative number of COVID-19 confirmed cases in Chinese cities (*x*) as of May 23, 2020. The main results from power law analyses appear in Tables 1-2 and Figure 5. As noted earlier, the requirement placed on sample size for credible power law analysis is a minimum of 50 observations (Clauset et al., 2009). This condition is well-satisfied here as the upper-tail sample contains 151 observations.^6^ The Hill and OLS estimates of the counter-cumulative parameter *γ* are around 0.80 and statistically different from zero. Given *m <* 0.80, the moments of the fitted power law distribution (including mean and variance) are generally non-convergent. The goodness-of-fit test of Gabaix and Ibragimov (2011) suggests we fail to reject the null hypothesis of power-law distributedness of COVID-19 cases in China.

**Table 1:** Power law parameter estimates and goodness-of-fit test.

|  | Estimate | Standard Error |
| --- | --- | --- |
| $\gamma^{Hill}$ | 0.808 | (0.066) |
| $\gamma^{OLS}$ | 0.762 | (0.087) |
| $x_{\min}$ | 24 | |
| Observations ($x > x_{\min}$) | 151 | |
| Observations (total) | 339 |  |
| *The Gabaix and Ibragimov goodness-of-fit test* |  |  |
| Goodness of fit test statistic | 0.013 |  |
| Goodness of fit threshold | 0.112 |  |

*Note:* Estimation is based on upper-tail observations $x > x_{\min}$ as of May 23, 2020, where $x_{\min}$ is determined based on the minimization of the KS statistic. For the Gabaix and Ibragimov \(2011\) test, the null hypothesis that COVID-19 confirmed cases is distributed according to a power law is rejected if a goodness-of-fit statistic is greater than a corresponding threshold value. Clauset et al. \(2009\) recommend to have at least 50 observations for accurate power law analysis, a condition satisfied here.

**Table 2:** Model fit measures of competing distributions.

|  | Log-likelihood | AIC | BIC | Likelihood ratio statistic | P-value |
| --- | --- | --- | --- | --- | --- |
| Power law | -845.545 | 1,693.089 | 1,696.106 |  |  |
| Exponential | -1,097.721 | 2,197.441 | 2,200.459 |  |  |
| Lognormal | -1,174.851 | 2,353.702 | 2,359.736 |  |  |
| Power law vs. exponential |  |  |  | 252.176 | 0.002 |
| Power law vs. lognormal |  |  |  | 329.306 | 0.000 |

*Note:* Estimation is based on upper-tail observations $x > x_{\min}$ as of May 23, 2020, where $x_{\min}$ is determined based on the minimization of the KS statistic. A positive value of the likelihood ratio statistic indicates that the power law is the better fitting distribution. A negative value indicates the alternative distribution fits the data more closely. P-values are calculated using the methods detailed in Clauset et al. \(2009\). The null hypothesis is there is no significant differences in likelihoods of the distributions tested.

**Figure 5:**
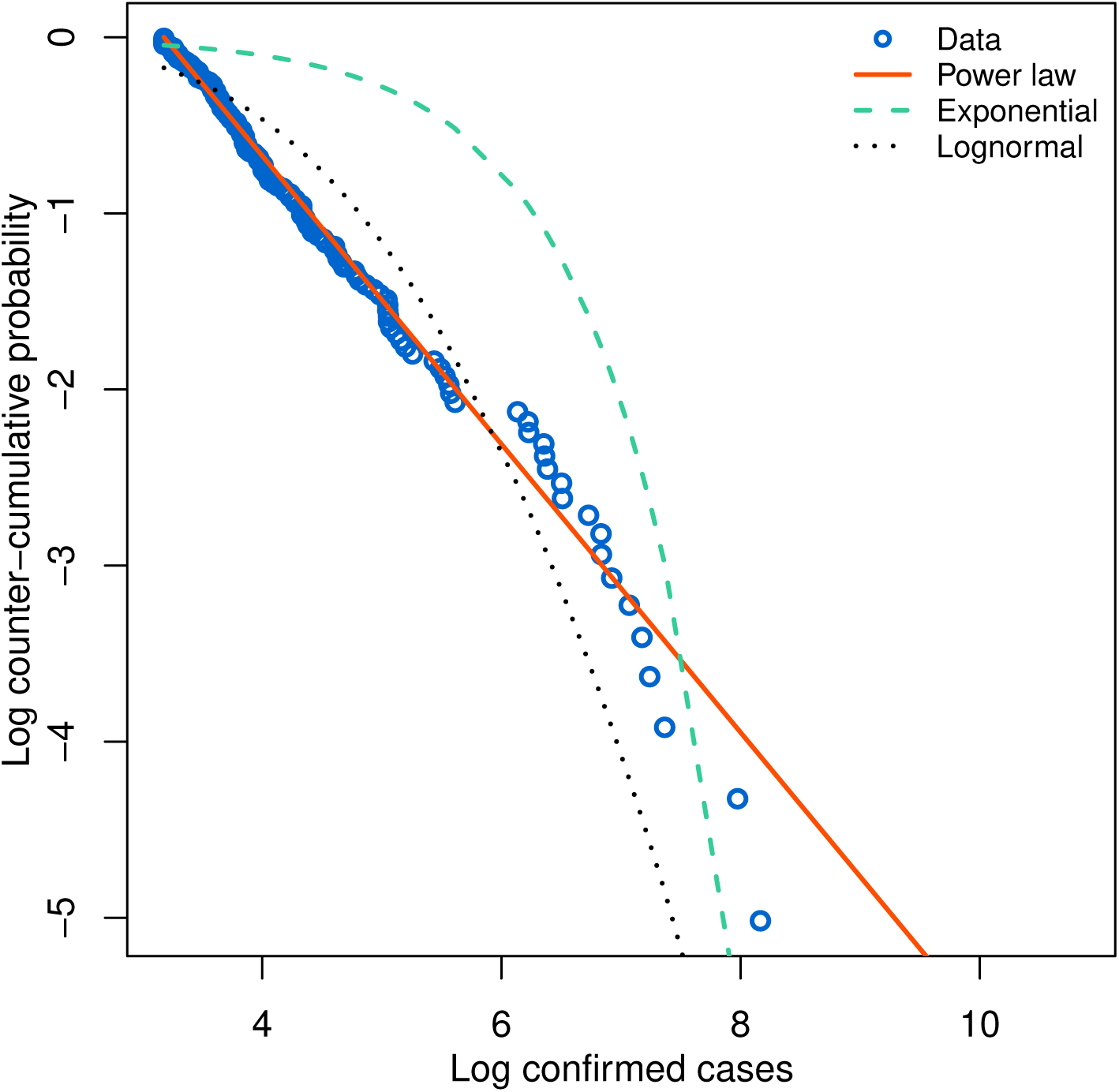
Plot of empirical and fitted log counter-cumulative probability and log COVID-19 confirmed cases. Estimation is based on upper-tail observations *x > x*_min_ as of May 23, 2020, where *x*_min_ is determined based on the minimization of the KS statistic.

Figure 5 depicts the power law and competing heavy-tailed distributions’ fits to the data. It is clear that the power law distribution generally fits the data better than the rivaling distributions, particularly in the lower to mid quantiles of the upper tail, where the observed data forms a distinct linear pattern. The power law slightly overestimates the frequency of the largest cases in the extreme upper tail (after log confirmed cases of about 7.8), where the fitted distribution decays relatively slowly. The fits of competing distributions— the lognormal and exponential—noticeably deviate from the empirical data throughout the domain. Formal goodness-of-fit measures in Table 2 provide further support in this regard. As is evident from AIC and BIC, as well as large positive likelihood ratio statistics, the power law distribution markedly outperforms both the lognormal and exponential distribution in fitting COVID-19 cases in China, which is in line with our observations from Figure 5. Therefore, we reject both the lognormal (and, by extension, the normal) and exponential as an adequate specification for COVID-19 cases in Chinese cities.

In order to verify the sensitivity of our findings to the (data-driven) choice of *x*_min_, we next perform power-law estimation and diagnostics with alternative candidates for *x*_min_. Figure 6 presents our results, where, as reflected by the horizontal axes, we treat each observation in the sample as a potential choice for *x*_min_. In panels (a) and (b), we plot the Hill and OLS estimates, respectively, of power law parameter *γ* as a function of *x*_min_. This amounts to what is commonly known as a Hill plot, which is a visual approach to choosing *x*_min_ (Clauset et al., 2009). Given the estimates of *γ* appear roughly stable beyond the data-driven choice of *x*_min_ (i.e., the vertical dashed line), this suggests that the data-driven choice of *x*_min_ is also plausible under the visual selection technique of *x*_min_. Furthermore, the goodness-of-fit tests in panels (c) and (d) establish that for *any* choice of *x*_min_: (i) we consistently fail to reject the null hypothesis of Gabaix and Ibragimov (2011) goodness-of-fit test, and (ii) the power law fit emerges superior according to BIC (and AIC, which is omitted in the interest of space). This illustrates the robustness of our observation that the distribution of COVID-19 cases in China is consistent with the characterization of power laws.

**Figure 6:**
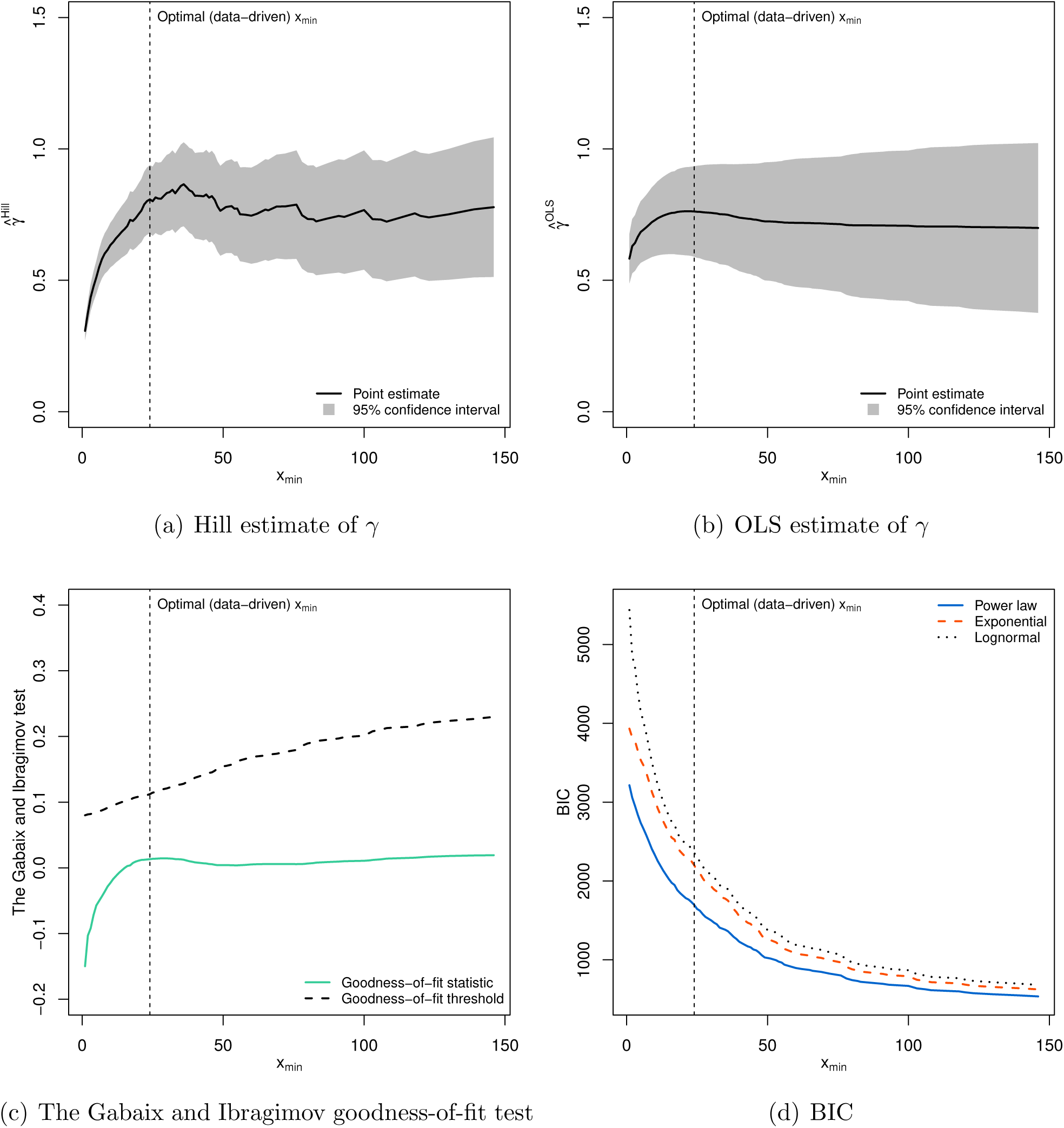
Sensitivity analysis with the choice of *x*_min_. Estimation for each given choice of *x*_min_ is based on upper-tail observations *x > x*_min_ as of May 23, 2020. The vertical (dashed) line indicates the position of optimal (data-driven) choice *x*_min_. For the Gabaix and Ibragimov (2011) test, the null hypothesis that COVID-19 confirmed cases is distributed according to a power law is rejected if a goodness-of-fit statistic is greater than a corresponding threshold value.

Lastly, we conduct a power law analysis through time to show a development, and possible trend, in power law exponent *γ*. This exercise also helps us test the robustness of our main finding to temporal variations and idiosyncrasies in the sample of COVID-19 cases in China. We fit the model to each day between January 15, 2020 and May 23, 2020, with *x*_min_ identified for each date using the method detailed in Section 3.1. According to panels (a) and (b) of Figure 7, the estimated value of power law parameter *γ* grows rapidly during the early days of the pandemic, when the virus was spreading most intensely, with the estimates of *γ* hovering around unity between late January and early February. By mid-February, the COVID-19 situation in China largely stabilized, which explains the trend forming around *γ* ≃ 0.8 in the period thereafter. Of particular interest for our subsequent analysis is the observation that *γ* ≃ 1 in the early phases of the outbreak. This indicates the existence of a *finite* mean for the distribution of COVID-19 cases in China during that time frame. Moreover, the diagnostic tests in panels (c) and (d) of Figure 7 indicate that the power law fit to COVID-19 cases in Chinese cities is remarkably robust over the first wave of the pandemic: (i) we consistently fail to reject the null hypothesis of Gabaix and Ibragimov (2011) goodness-of-fit test, and (ii) the power law fit remains superior according to BIC (and AIC, which is omitted in the interest of space). This shows that our main results reported in Tables 1-2 and Figure 5 are not driven by the nature of a sample used in that analysis, but rather power law is a genuine phenomenon in this case. Granted that the number of confirmed cases is affected by the underlying infective process as well as detection process, the power law analysis through time also establishes the robustness of our main finding to any potential changes in the detection process of COVID-19 cases over the course of the first wave of the pandemic.

**Figure 7:**
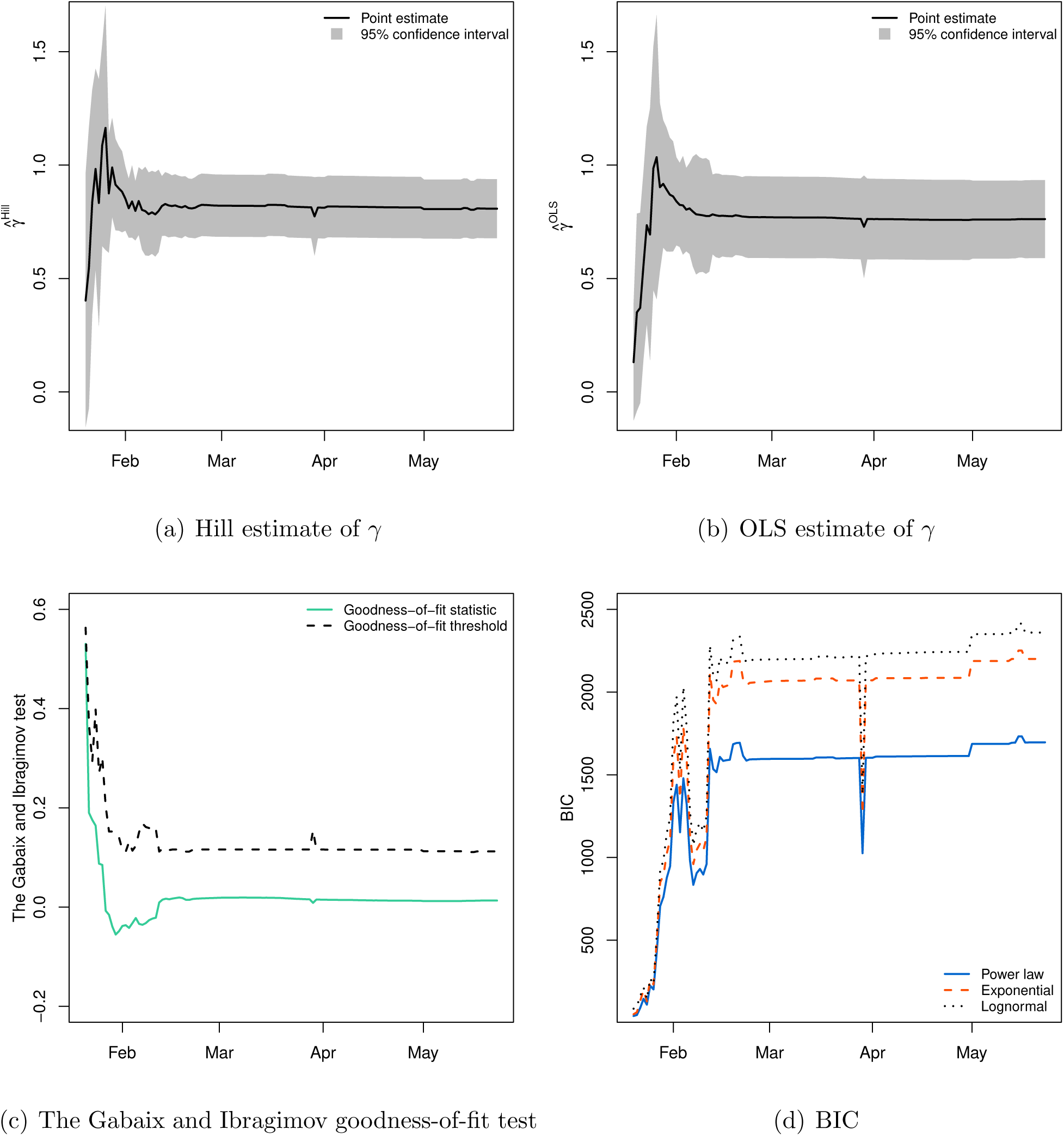
Power law analysis over the first wave of the pandemic. Estimation is based on upper-tail observations *x > x*_min_ on each day, where *x*_min_ for each date is determined based on the minimization of the KS statistic. For the Gabaix and Ibragimov (2011) test, the null hypothesis that COVID-19 confirmed cases is distributed according to a power law is rejected if a goodness-of-fit statistic is greater than a corresponding threshold value.

It is worth mentioning that the distribution in (1) is a representative from the class of regularly varying distributions (i.e., general power law). The power law estimation framework presented in this study is consistent under the assumption that a given data comes from a pure power law, which forms a small subset of general power laws. On the other hand, the estimation framework reviewed in Voitalov et al. (2019) is shown to be consistent under the more general assumption that the data comes from any impure power law. Indeed, there are more flexible forms of the Pareto distribution—often with an extra parameter and/or of mixture form—that allow for capturing extreme upper-tail probabilities more closely. Such generalized, or, composite, distributions have been shown to improve upon the benchmark power law distribution in fitting empirical data (see, for instance, Giesen et al., 2010, Patel and Schoenberg, 2011, Ioannides and Skouras, 2013, Luckstead and Devadoss, 2017, and Nigai, 2017). The main goal of the present study is to determine whether a power law generally approximates the *upper-tail* COVID-19 cases in China, which has an implication for understanding tail risk properties of COVID-19, and not the investigation of various distributions within the power law family, which is beyond the scope of this research. Our estimation results and diagnostic tests provide empirical evidence that the right-tail of COVID-19 cases in Chinese cities is well-characterized by the power law (Pareto) distribution.

## 4 Explaining a power law in COVID-19 cases in China

There are different mechanisms proposed in the literature that can generate power laws.^7^ In this section, we explore whether a growth model involving Gibrat’s law (Gibrat, 1931) can potentially explain the emergence of the observed power-law behavior in COVID-19 cases in China. We focus on Gibrat’s law specifically granted a random multiplicative growth is the prevalent attribute of models explaining the genesis of power laws (Reed, 2001; Gabaix, 1999, 2009).

### 4.1 A link between power law and Gibrat’s law

Suppose *S*_*it*_ is the size of a stochastic process of interest for unit *i* at time *t*. For instance, COVID-19 cases in city *i* up to day *t*. According to Gibrat’s law, the size of the process (at least in the upper tail) exhibits random multiplicative growth, evolving as

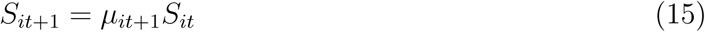

over time, where *μ*_*it*+1_ is independently and identically distributed (i.i.d.) random variable with an associated PDF of *f* (*μ*). Hence, random growth factor *μ*_*it*+1_ = *S*_*it*+1_*/S*_*it*_ is independent of the current size *S*_*it*_, which is commonly known as Gibrat’s law of proportionate growth. Gibrat’s law alone is not sufficient to give rise to a power law. In fact, it leads to the lognormal distribution, which was noted by Gibrat (1931) himself early on.^8^ Gabaix (1999) showed that power law can arise from Gibrat’s law with an auxiliary assumption, a sketch of which we provide below.

Let *G*_*t*_(*s*) = Prob(*S*_*t*_ *> s*) be the counter-CDF of *S*_*t*_. Substituting (15) into the counter-CDF, the equation of motion for *G*_*t*+1_(*s*) boils down to

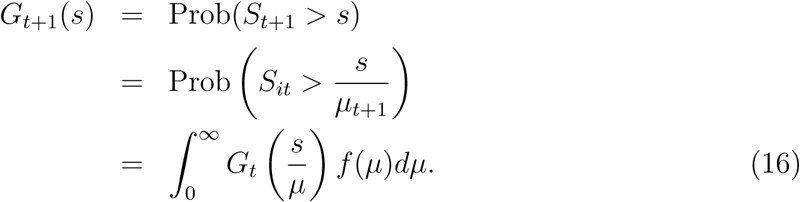

If there is a steady state process *G*_*t*_ = *G*, then

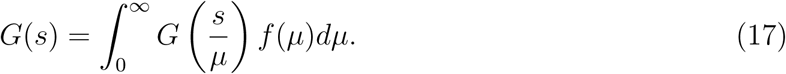

The mechanism ensuring that power law distribution is the (only) suitable steady state distribution in (17) is if *S*_*t*_ has lower reflecting barrier *S*_min_, i.e., the minimal size of the process, such that *S*_*t*_ *> S*_min_ (Gabaix, 1999, Proposition 1). In this case, 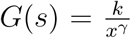, from (8). Thus, Gibrat’s law combined with a lower bound on *S*_*t*_ can plausibly yield power law distribution.

The standard theory of proportionate random growth is valid if the mean of the stationary distribution is finite (Gabaix, 1999, 2009). Our analysis in Section 3.3 uncovers a power law in COVID-19 cases (as of May 23, 2020) with no finite mean, which seemingly indicates that the specific power law for COVID-19 cases in China cannot be generated by the standard model for Gibrat’s law of proportionate growth. But, as our temporal power-law analysis has demonstrated (see Figure 7), the fitted power-law distribution does have a finite mean during the early phases of the outbreak (as *γ* ≃ 1), when the number of confirmed cases was rising. This implies a proportionate random growth model can offer a possible explanation for the emergence of a power law in the distribution of COVID-19 cases in the early phases of the outbreak.^9^ In the later phases of the outbreak, as the number of cumulative cases starts to stabilize, a proportionate random growth model becomes less and less plausible, as E(*μ*_*it*_) = 1 in (15). This is anticipated as, after all, Gibrat’s law of proportionate growth is a model of growth. Therefore, we formally test for proportionate random growth at micro-level by analyzing growth rates of COVID-19 cases in Chinese cities during the early days of the pandemic.

### 4.2 Testing for Gibrat’s law

For empirical purposes, we consider a continuous time representation of Gibrat’s law, given by geometric Brownian motion

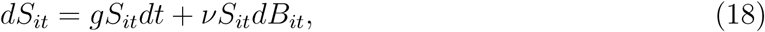

where *g* is the expected growth rate, *ν* > 0 is the volatility, and *B*_*it*_ is a standard Brownian motion that is i.i.d. across cross-sectional units. Applying Ito’s lemma to (18) yields

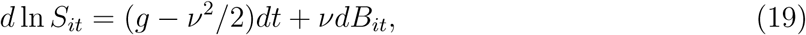

meaning the cross-sectional distribution of *S*_*it*_, with the initial size of *S*_*i*0_, is lognormal

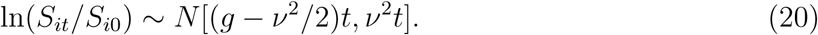

Equation (19), along with Proposition 1 in Gabaix (1999), suggests that growth rates under Gibrat’s law can be described by a random walk process of the form (Sutton, 1997; Eeckhout, 2004; Gabaix, 2009)

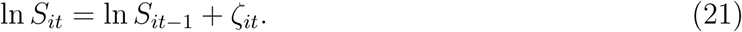

Setting the random growth component *ζ*_*it*_ = *φ*_*i*_+*ξ*_*it*_, where *φ*_*i*_ is the effect of unit-wide factors and *ξ*_*it*_ is an i.i.d. random effect, produces a random walk with drift. A standard method to test for Gibrat’s law is through estimation of the following cross-sectional regression equation

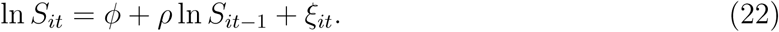

In (22), *ρ* is the parameter of interest, with *ρ* ≃ 1 providing statistical evidence that the growth process of *S*_*t*_ adheres to Gibrat’s law.

An alternative approach for testing for proportionate random growth is through estimation of the cross-sectional regression equation of the form (Beare and Toda, 2020)

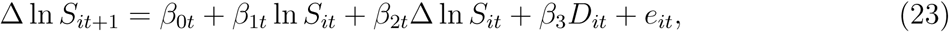

where Δ is the difference operator, Δ ln *S*_*it*+1_ is the COVID-19 growth rate in city *i* between day *t* and *t* + 1, Δ ln *S*_*it*_ is the COVID-19 growth rate in city *i* between day *t* − 1 and *t, D*_*it*_ is the number of days between day *t* and the day of the first COVID-19 case in city *i*, and *e*_*it*_ is an i.i.d. error term. The parameters of interest are *β*_1*t*_, *β*_2*t*_, *β*_3*t*_, with *β*_1*t*_ ≃ 0, *β*_2*t*_ ≃ 0, *β*_3*t*_ ≃ 0 providing empirical evidence for the presence of Gibrat’s law. A distinctive feature of equation (23) is the inclusion of age distribution—days since outbreak for each city—in addition to the growth rate. Obtaining age distribution has traditionally been cumbersome in power law analysis (e.g., of cities). Fortunately, our data conveniently affords us this variable as we observe the entire timeline of the evolution of COVID-19 across Chinese cities.

### 4.3 Application

As discussed in Section 4.1, a proportionate random growth model can potentially explain a power law in the data during the early phases of the pandemic. We thus apply the methods reviewed in Section 4.2 to each day between January 23, 2020, and February 2, 2020 (inclusive). The reason for choosing these dates is twofold. First, at least 30 cities had a nonzero number of cumulative cases (*S*_*it*_ *>* 0) during that period (see Figure 2). Second, the estimated power law exponent is *γ* ≃ 1 between late January and early February (see Figure 7), which is a condition necessary for the validity of a proportionate random growth process, as noted above.

Figure 8 shows the estimation results for *ρ*_*t*_ in equation (22) for *t* = Jan 23, …, Feb 2. Clearly, the estimates of *ρ*_*t*_ are statistically indistinguishable from unity (*ρ*_*t*_ ≃ 1), which confirms the random growth model predicated by Gibrat’s law. The 95% confidence interval shrinks moving left to right, which can be attributed to increasing sample size (i.e., increasing number of cities with confirmed cases) over time (see Figure 1).

**Figure 8:**
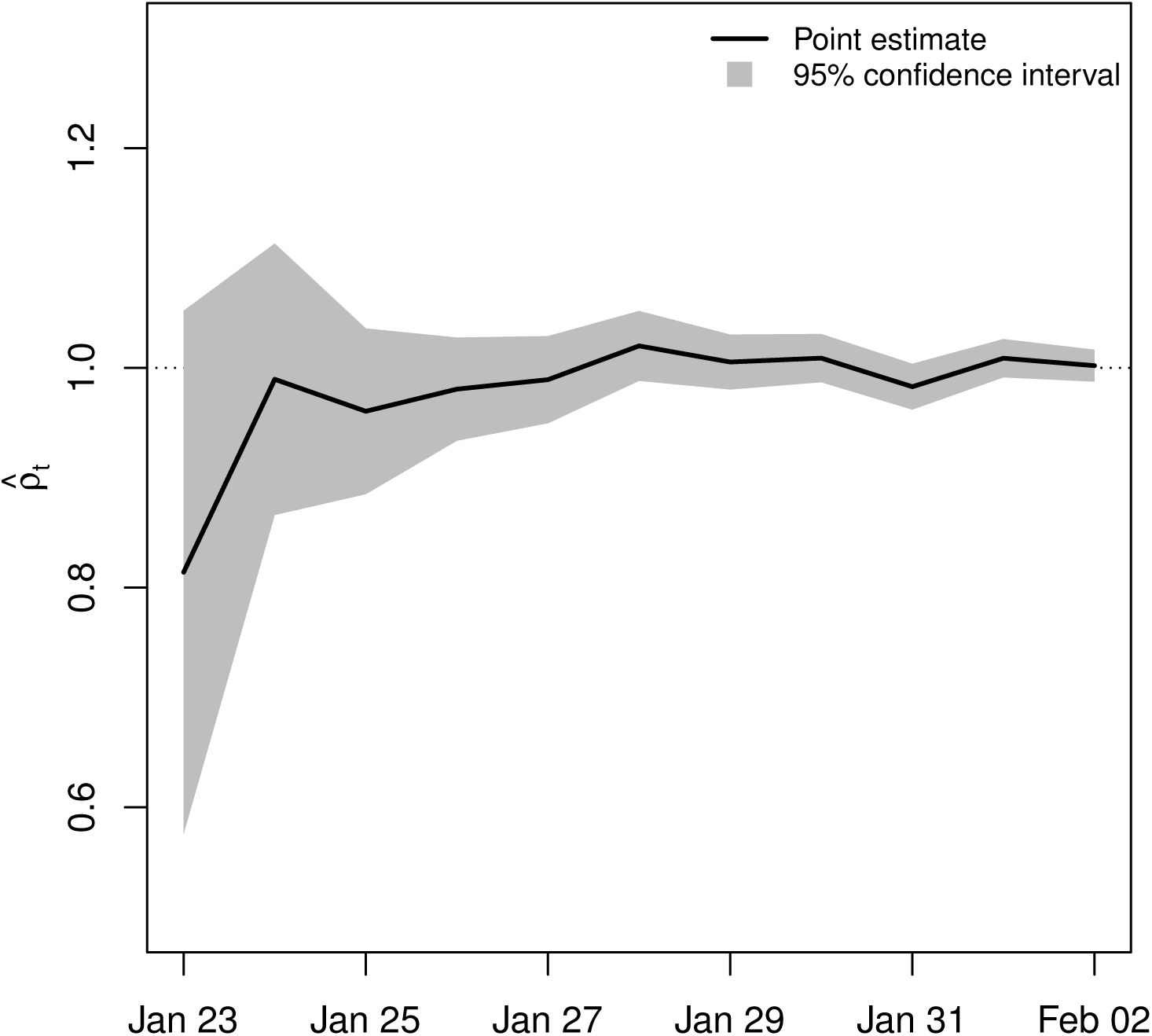
Estimates of *ρ*_*t*_ in equation (22) between January 23, 2020, and February 2, 2020, with 95% confidence bands. *ρ*_*t*_ ≃ 1 provides empirical evidence for Gibrat’s law.

Figure 9 reports the estimation results for *β*_0*t*_, *β*_1*t*_, *β*_2*t*_, *β*_3*t*_ in equation (23) for *t* = Jan 23, …, Feb 2. Panels (b)–(d) contain the estimates for *β*_1*t*_, *β*_2*t*_, *β*_3*t*_, which are of primary interest here. It is apparent that these estimates are mostly equal to zero (*β*_1*t*_ ≃ 0, *β*_2*t*_ ≃ 0, *β*_3*t*_ ≃ 0), which indicates the growth rate between days *t* and *t* + 1 does not depend on the number of cases on day *t*, nor on the growth rate between days *t* − 1 and *t*, nor on the number of days since the first confirmed case. This provides strong evidence for the existence of Gibrat’s law for COVID-19 cases in Chinese cities during the period under consideration. The estimates of *β*_0*t*_ in panel (a) show that the expected growth rate of confirmed cases was indeed positive between January 23 and February 2, with a general downward trend.

**Figure 9:**
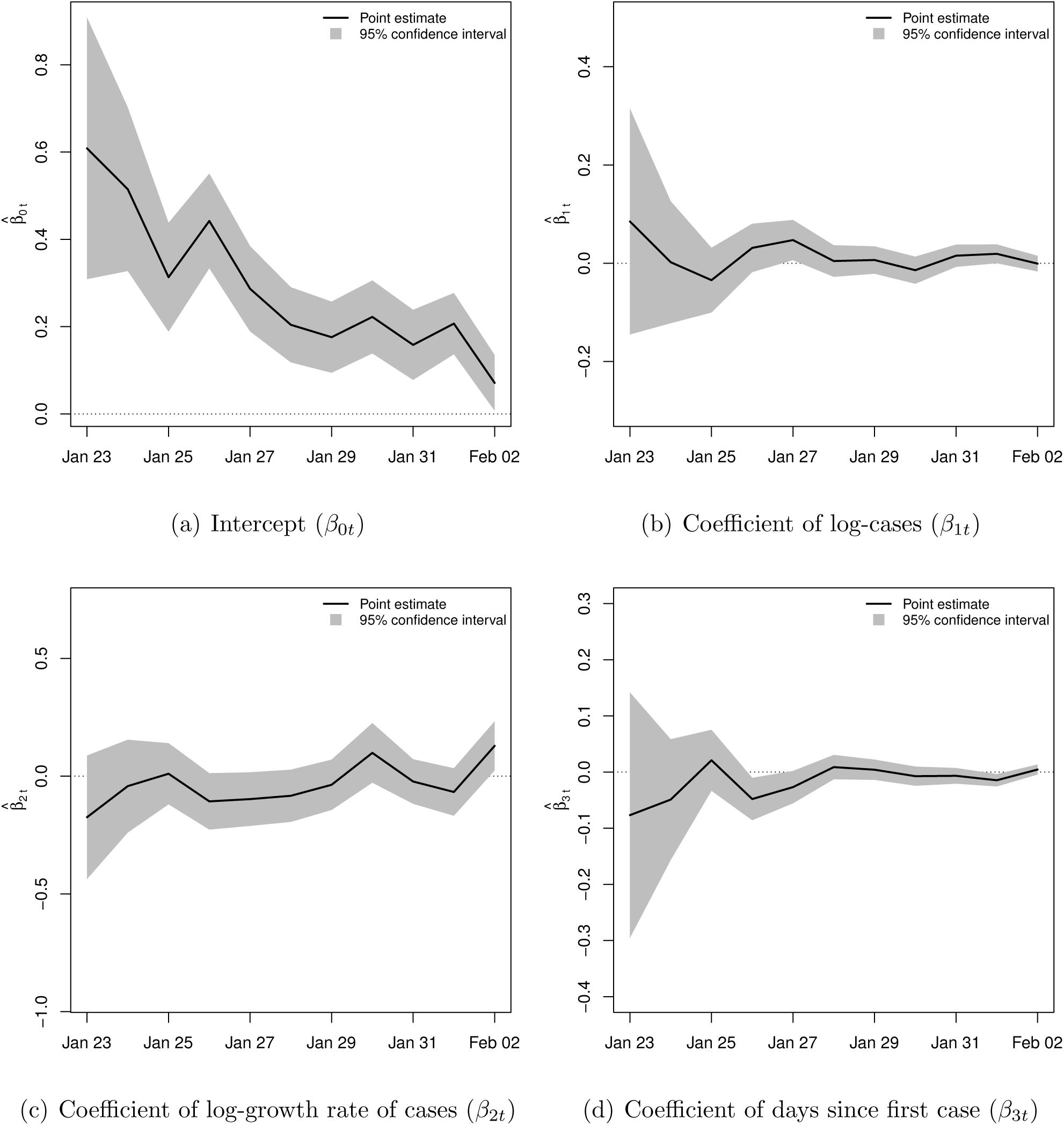
Estimates of *β*_0*t*_, *β*_1*t*_, *β*_2*t*_, *β*_3*t*_ in equation (23) between January 23, 2020, and February 2, 2020, with 95% confidence bands. The parameters of interest are *β*_1*t*_, *β*_2*t*_, *β*_3*t*_, with *β*_1*t*_ ≃ 0, *β*_2*t*_ ≃ 0, *β*_3*t*_ ≃ 0 providing empirical evidence for Gibrat’s law.

In summary, confirmation of Gibrat’s law provides support for a proportionate random growth model as a plausible power-law generating mechanism for COVID-19 cases in China in the early phases of the outbreak, when the number of cases was still rising. In addition, confirmation of Gibrat’s law implies that COVID-19 cases in Chinese cities grew proportionately during the early phases of the pandemic (when the number of cases was rising), with the underlying stochastic process remaining the same for all cities. This means each city’s COVID-19 growth rate was drawn randomly and independently of its size. In the later phases of the outbreak, as the number of cumulative cases starts to stabilize, a proportionate random growth model becomes less plausible, as E(*μ*_*it*_) = 1 in (15), hence the underlying stochastic process is no longer the same for all cities.

## 5 Conclusion

The dynamics of the novel coronavirus pandemic are complex and determined by a myriad of factors, which are yet to be fully understood. In spite of the apparent chaotic evolution of the pandemic, surprising regularities can still be observed in the size distribution and growth process of COVID-19 cases. In this article, we examined the distribution of the novel coronavirus cases in China—the original epicenter of the ongoing pandemic. We presented empirical evidence for a power law distribution for the upper tail of the number of COVID-19 cases in Chinese cities. The power law fit is robust to different estimation methods, passes rigorous diagnostic tests, and fits the data better than a number of natural alternatives. As such, the power law surmounts the statistical hurdle.

The implications of the power law fit are that (i) the number of COVID-19 cases in Chinese cities is heavy-tailed and disperse; (ii) mean and variance are generally not finite, so that average number of COVID-19 cases is problematic to talk about; and (iii) COVID-19 cases are concentrated within a few cities that account for a disproportionately large amount of infections. Admittedly, there may always be a distribution that fits the data better than a power law granted there are virtually an infinite number of distributions. What we showed in this study is that the power law distribution is able to capture the upper tail of the data, and better than several ‘go-to’ distributions. This has important implications for empirical work as, for reliable analysis of the effects of COVID-19 pandemic, using econometrically justified and robust methods that account for potential heavy-tailedness and tail risk property is in order.

## Data Availability

The data used in this research project is publicly available. We provide online links to the data in the project.

## Footnotes

1 For a detailed review of power laws, see Reed (2001), Newman (2005), Sornette (2006) and Gabaix (2009, 2016). Notably, the power law distribution with exponent *γ* ≃ 1 generates Zipf’s law (Zipf, 1949).

2 For a detailed review of Gibrat’s law, see Sutton (1997).

3 In particular, Gibrat’s law has been shown to explain the growth process of consumption (Battistin et al., 2009), firms (Luttmer, 2007), farms (Clark et al., 1992), cities (Ioannides and Overman, 2003; Eeck-hout, 2004), countries (Rose, 2006; González-Val and Sanso-Navarro, 2010), carbon dioxide (CO_2_) emissions (Ahundjanov and Akhundjanov, 2019), and bird population (Keitt and Stanley, 1998), among others.

4 We use the 2020 population data for Chinese cities collected from United Nations (2018).

5 For other properties of the likelihood ratio statistic, including its advantage over other goodness-of-fit measures and the derivation of its p-value, see Clauset et al. (2009).

6 The KS estimate of *x*_min_ is 24, as reported in Table 1. Consequently, 151 observations out of 339, which satisfy *x* ≥ 24, are used in power law analysis in this case.

7 For a detailed review, see Reed (2001), Mitzenmacher (2004), Newman (2005), and Gabaix (2016).

8 Interestingly, many examples used by Gibrat (1931) have recently been shown to actually follow a Pareto-type distribution rather than the lognormal (Akhundjanov and Toda, 2020).

9 Similar observation was made by Beare and Toda (2020) for COVID-19 cases in US counties.

